# Severity detection for the coronavirus disease 2019 (COVID-19) patients using a machine learning model based on the blood and urine tests

**DOI:** 10.1101/2020.07.27.20044990

**Authors:** Haochen Yao, Nan Zhang, Ruochi Zhang, Meiyu Duan, Tianqi Xie, Jiahui Pan, Ejun Peng, Juanjuan Huang, Yingli Zhang, Xiaoming Xu, Hong Xu, Fengfeng Zhou, Guoqing Wang

**Author notes:** Correspondence: Guoqing Wang,; Fengfeng Zhou,; Hong Xu. These authors have contributed equally to this work (Haochen Yao, Nan Zhang, Ruochi Zhang).

## Abstract

The recent outbreak of the coronavirus disease-2019 (COVID-19) caused serious challenges to the human society in China and across the world. COVID-19 induced pneumonia in human hosts and carried a highly inter-person contagiousness. The COVID-19 patients may carry severe symptoms, and some of them may even die of major organ failures. This study utilized the machine learning algorithms to build the COVID-19 severeness detection model. Support vector machine (SVM) demonstrated a promising detection accuracy after 32 features were detected to be significantly associated with the COVID-19 severeness. These 32 features were further screened for inter-feature redundancies. The final SVM model was trained using 28 features and achieved the overall accuracy 0.8148. This work may facilitate the risk estimation of whether the COVID-19 patients would develop the severe symptoms. The 28 COVID-19 severeness associated biomarkers may also be investigated for their underlining mechanisms how they were involved in the COVID-19 infections.

## INTRODUCTION

Multiple cases of pneumonia patients were linked to the coronavirus disease-2019 (COVID-19) occurred in December 2019 [1]. The virus 2019-nCoV demonstrated a substantial capability of inter-human transmissions [2] and has rapidly spread around the world, in particular South Korea and Japan [3]. Patients infected with COVID-19 had significantly varied symptoms and their outcomes ranged from mild to death, and the mortality rate was approximately 4.3% [4]. It is necessary to mention that 61.5% of the COVID-19 pneumonia patients with critical symptoms died within 28 days after admission [5]. The discrimination of severely ill patients with COVID-19 from those with mild symptoms may help understand the individualized variations of the COVID-19 prognosis. The knowledge may also facilitate the establishing of early diagnosis of the COVID-19 severeness.

The diagnosis of COVID-19 heavily relies on the epidemiological features, clinical characteristics, imaging findings, and nucleic acid screening [6], etc. The delivery of the diagnosis result by these technologies was time consuming and error prone [7]. Multiple types of clinical data were collected for a patient with COVID-19 infection and they were manually integrated by the clinicians to make the diagnosis decisions. The stochastic transmission model was also used to investigate how the COVID-19 transmitted locally and globally [8]. Machine learning algorithms were widely used to integrate the heterogeneous biomedical data sources for the diagnosis decision [9,10]. So they may also be utilized to produce more delicate prediction models for the severeness diagnosis of the COVID-19 patients. The biomarkers used for an accurate diagnosis model of patients with COVID-19 may serve as the drug targets for this global infectious disease.

This study investigated the detection of severely ill patients with COVID-19 from those with mild symptoms using the clinical information and the blood/urine test data. The clinical information consisted of age, sex, body temperature, heart rate, respiratory rate and blood pressure. The blood/urine tests may be carried out using the technically-easy and cost-efficient procedures. An accurate severeness detection model of the patients with COVID-19 based on those features above may improve the prognosis of this disease in large scale clinical practices. The following sections will firstly describe the data collection and modeling methods, and then utilized the popular machine learning algorithms to build the best severeness detection model.

## MATERIALS AND METHODS

### Data collection

This study recruited 137 clinically confirmed cases of COVID-19, which were collected from the Tongji Hospital Affiliated to Huazhong University of Science and Technology. Patients were hospitalized from January 18, 2020, to February 13, 2020. The cohort consisted of 17 mild cases, 45 moderate ones and 75 severely ill patients. 21 of the severe cases eventually died. This study investigated the binary classification problem between 75 severe/deceased cases and 62 mild/moderate ones. Each participant was regarded as a sample in this study. This study was approved by the Ethics Commission of the First Hospital of Jilin University(2020-236). With informed consent was waived for this emerging infectious disease.

Patient information including age, sex, body temperature, heart rate, respiratory rate, blood pressure and the blood/urine tests data. Each clinically-obtained value was regarded as a feature in this study. In summary, each sample has 100 features, consisting of 8 clinical, 76 blood test and 16 urine test values.

### Data pre-processing

The missing entries were filled in the following procedure. We assumed a missing entry to be within the normal range and filled this entry with the median of that normal range. If there is no normal range for a missing entry, we filled it with zero (0). The samples were randomly split into 80% as training and 20% as test datasets in a stratified fashion. Features in continuous values were normalized by the values in the training dataset. The categorial features were encoded by the one-hot strategy.

### Feature selection

The principle of Occam’s razor suggested that a model using fewer features was preferred over a complicated model with a similar prediction performance [11]. Feature selection algorithms may be utilized to remove those unrelated features [12] and may usually increase the model prediction performances [13,14].

The student t-test (abbreviated as T-test) is a filter algorithm and it evaluates the statistical association of each feature with the disease severeness of a sample. The features with the T-test calculated Pvalues below 0.05 were usually considered to be statistically significantly associated with the disease severeness [15,16].

### Prediction algorithms

This study evaluated several classification algorithms to build the prediction models of the severely ill patients with COVID-19. The predictive logistic regression (LR) model is a regression analysis for the dataset with the binary dependent variable, i.e., the class label [17]. LR has been widely used to build clinical decision models [18,19]. A probability is calculated by LR to describe whether the sample belongs to a class and a threshold for the probability is usually utilized to make the predictive decision. LR firstly calculates the log-odds l=log_b_[p/(1-p)]=β_0_+β_1_x+…+β_n_x, and the probability p=1/[1+b^-(β0+β1x+…+βnx)^], where β_i_ is the model parameter.

Support vector machine (SVM) is a supervised machine learning algorithm that may accomplish both classification and regression tasks [20]. SVM tries to find a hyperplane to separate data by the highest margin. The learning strategy of SVM is spacing maximization, which can be formalized as a problem of solving convex quadratic programming [21]. This algorithm has been widely used to build the prediction models using the data of blood test [22,23] and urine test [24,25].

Random forest (RF) is an ensemble algorithm that summarizes the prediction results of multiple tree-based classifiers [26]. RF may improve the model performances and avoid over-fitting by averaging the results of models trained over various sub-samples of the dataset. Its model complexity renders itself computation-intensive and RF runs slower than many prediction algorithms. RF is another popular algorithm for building the prediction models using the clinical data [27,28].

K nearest neighbor (KNN) is an instance-based learning algorithm and summarizes the prediction based on the class labels of the query sample’s k nearest neighbors [29]. KNN simply assigns the query sample with the class label of its majority nearest neighbors. And its prediction performance heavily relies on the definition of the inter-sample distances. Nicholas Schaub, et al., demonstrated that selecting the best biomarkers may be essential to improve the KNN models [30].

The boosting-based algorithm AdaBoost iteratively trained weak learners and summarized these weak learners’ results into a weighted sum [31]. Multiple variants of Adaboost were proposed for recognizing human actions [32], diagnosing the dog hypoadrenocorticism [33], and predicting protein binding sites [34], etc.

The above algorithms are implemented using Python programming language (version 3.6) and Scikit-learn package (version 0.22).

### Prediction performance evaluation metrics

The binary classification model was evaluated using four classification performance metrics, as defined in the followings. The severely ill patients were regarded as positive samples and the other patients constituted the negative dataset. The number of correctly predicted positive samples was defined as true positive (TP), and the number of the other positive samples was false negative (FN). The true negative (TN) and the false positive (FP) were defined as the numbers of correctly and incorrectly predicted negative samples, respectively. So the overall accuracy Acc was defined as Acc=(TP+TN)/(TP+FN+TN+FP). The model’s sensitivity (Sn) and specificity (Sp) were defined as Sn=TP/(TP+FN) and Sp=TN/(TN+FP). The three metrics Acc, Sn and Sp measured the percentages of correctly predicted all, positive and negative samples, respectively. The Matthew’s Correlation Coefficient (MCC) described the overall correlation of the predicted and the real class labels, and MCC was defined as MCC=(TP×TN-TP×FN)/sqrt[(TP+FP) ×(TP+FN) ×(TN+FP) ×(TN+FN)], where sqrt() was the square root function [35,36].

Each model was randomly trained for twenty runs with different random seeds and the metric averaged accuracy aAcc=[Acc(1)+Acc(2)+…+Acc(20)]/20, where Acc(i) was the accuracy of the i^th^ model. The metric aAcc was used to find the best prediction model. The metrics aSn, aSp and aMCC were the averaged Sn, averaged Sp and averaged MCC over the twenty random runs.

### Ethics statement

This study was approved by the Ethics Commission of the First Hospital of Jilin University(2020-236). With informed consent was waived for this emerging infectious disease.

## RESULTS

### Baseline characteristics of the 2019-nCoV pneumonia participants

This study recruited 137 COVID-19 patients to build the detection model of severely ill (positive) samples against the patients with mild symptoms. All the 100 features were screened for their association with the class label, i.e., Positive or Negative. There were 8 clinical values, 76 blood test values and 16 urine test values, respectively. Thirty-two features achieved the T-test Pvalue<0.05, and were kept for further analysis in the following sections,as summarized in the Supplementary Table S1.

The feature of the patient’s age at diagnosis (Age) demonstrated a significant difference (Pvalue=1.75e-6) between the two groups of samples, and the severely ill patients were on average 13.5695 years older than the patients with mild symptoms. This supported the observation that patients aged around 65 years old tended to have more severe symptoms than those aged around 51 years old [37]. The sex also demonstrated severe-specific Pvalue=7.71e-5, suggesting that male patients were at higher risks of developing severe symptoms [38], as shown in Figure 1 (A).

**FIGURE 1.**
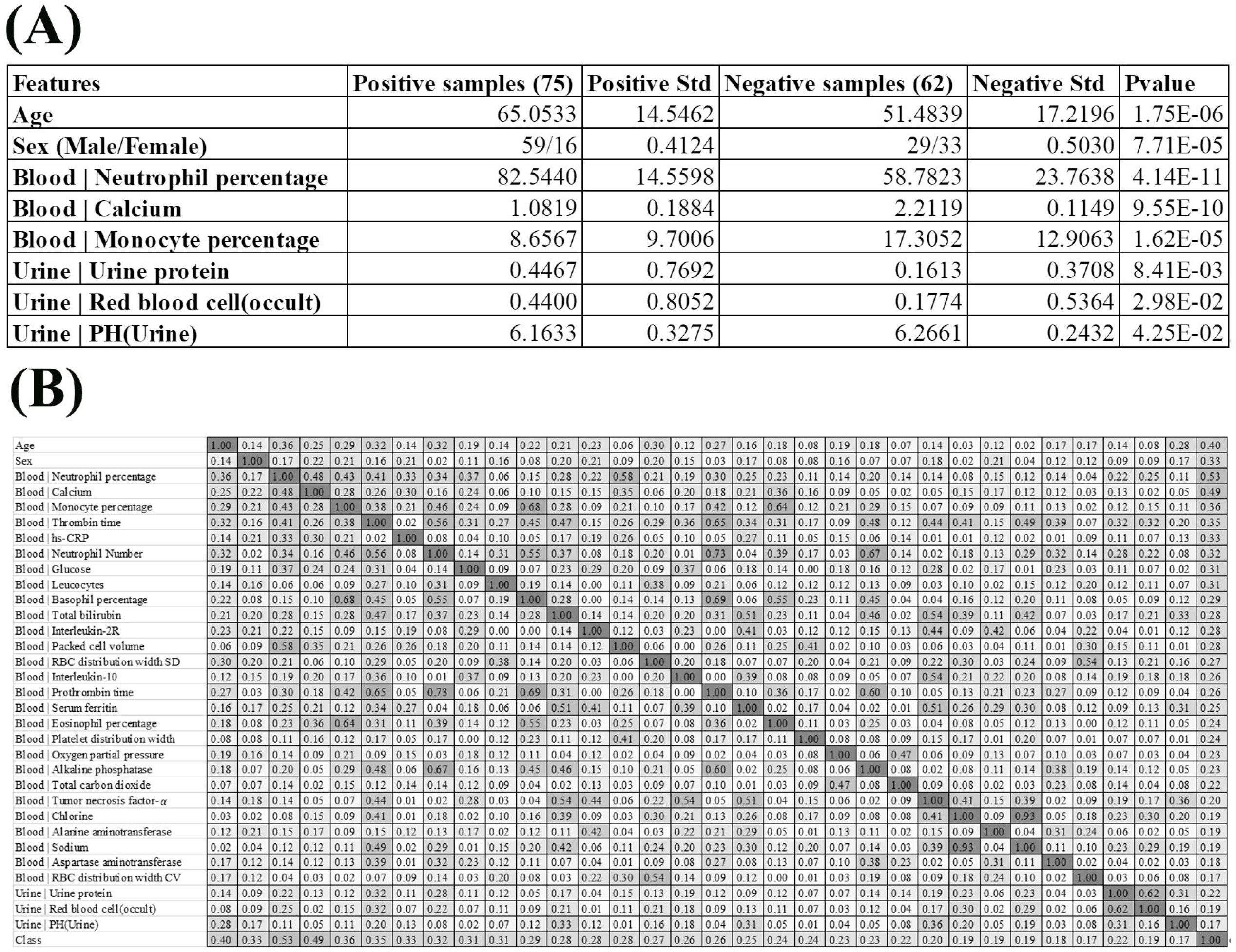
Baseline summary of the recruited cohort. **(A)**There were 75 positive and 62 negative samples, respectively. The columns “Positive Std” and “Negative Std” gave the standard deviations of the specific feature in each sample group. The last column “Pvalue” gave the T-test Pvalue of that specific feature between the two sample groups. A feature name starting with “Blood | “and “Urine | “was collected from the blood test and urine test, respectively. **(B)** The heatmap matrix of the inter-feature Pearson correlation coefficient (PCC) for all the features and the group value. The values ranged between 0.00 and 1.00, and the color was linearly rendered according the inter-feature PCC. The feature names starting with “Blood | “and “Urine | “were from the blood test and urine test, respectively.

We also summarized three blood test values and three urine test values with the most significant differences between the two groups of samples, as shown in Figure 1 (A). Overall, the blood test values demonstrated much more significant inter-group differences than the urine test values. The summary data suggested that the percentage of neutrophil cells was significantly enriched in the blood of the severely ill patients, with P values 4.14e-11. In addition, the serum calcium level and the monocyte percentage were also significantly lower in the severely ill patients than those mild ones.

Three urine test values demonstrated weak inter-group differential significances. The two values “Urine | Urine protein” and “Urine | Red blood cell(occult)” demonstrated the elevated levels in the severely ill patients with Pvalues 1.44e-2 and 2.83e-2, respectively. But their variations were very larger, which rendered neither of them as good disease severeness biomarkers. A minor decrease (0.1028) in the urine pH value (feature “Urine | PH(Urine)”) in the severely ill patients achieved the inter-group differential significance Pvalue 4.25e-2.

In the following sections. The detailed summary may be found in the Supplementary Table S1.

### Evaluation of feature correlations with the group labels

We firstly evaluated the correlation between the 32 features and the class label using Pearson Correlation Coefficient (PCC), as shown in Figure 1 (B). The PCC value ranges between -1 and 1. This study focused on the whether a feature was correlated with the class label. So the absolute value of PCC was calculated in Figure 1 (B).

Some features showed strong correlations with the 2019-nCoV pneumonia severeness, which was the class label. The feature “Blood | Neutrophil percentage” demonstrated the largest PCC=0.53 with the disease severeness (class label). This provided another piece of evidence that the neutrophil cell percentage was positively correlated with the 2019-nCoV severeness. Another feature “Blood | Calcium” achieved the second-best PCC=0.49. The age at diagnosis (feature Age) achieved the third-best PCC=0.40 with the class label, suggesting that the elder patients were under higher risks of developing severe symptoms.

Figure 1 (B) suggested that some of the 32 features were highly correlated with the class label and they may facilitate the training of a reasonably-accurate detection model for the 2019-nCoV pneumonia severeness. The existence of high inter-feature correlations suggested that some redundant features may need to be removed to further improve the detection model.

### Comparison of different prediction algorithms

Five prediction algorithms were evaluated for their detection performances using their default parameters on all the 98 features of the 2019-nCoV pneumonia patients, as shown in Figure 2. Firstly, all the five prediction algorithms achieved at least 0.7130 in Acc on all the 32 features, suggesting that the severely ill COVID-19 patients may have severeness-specific patterns. The prediction algorithm SVM achieved the best prediction accuracy Acc=0.7926 and its standard deviation in Acc was only 0.0715. SVM achieved the sensitivity Sn=0.7666 much better than the specificity Sp=0.6993. The prediction sensitivity was the detection accuracy of the positive samples, i.e., the severely ill patients. So the following sections used the prediction algorithm SVM as the default predictor and the prediction model was further refined by optimizing the SVM parameters and selecting the best features.

**FIGURE 2.**
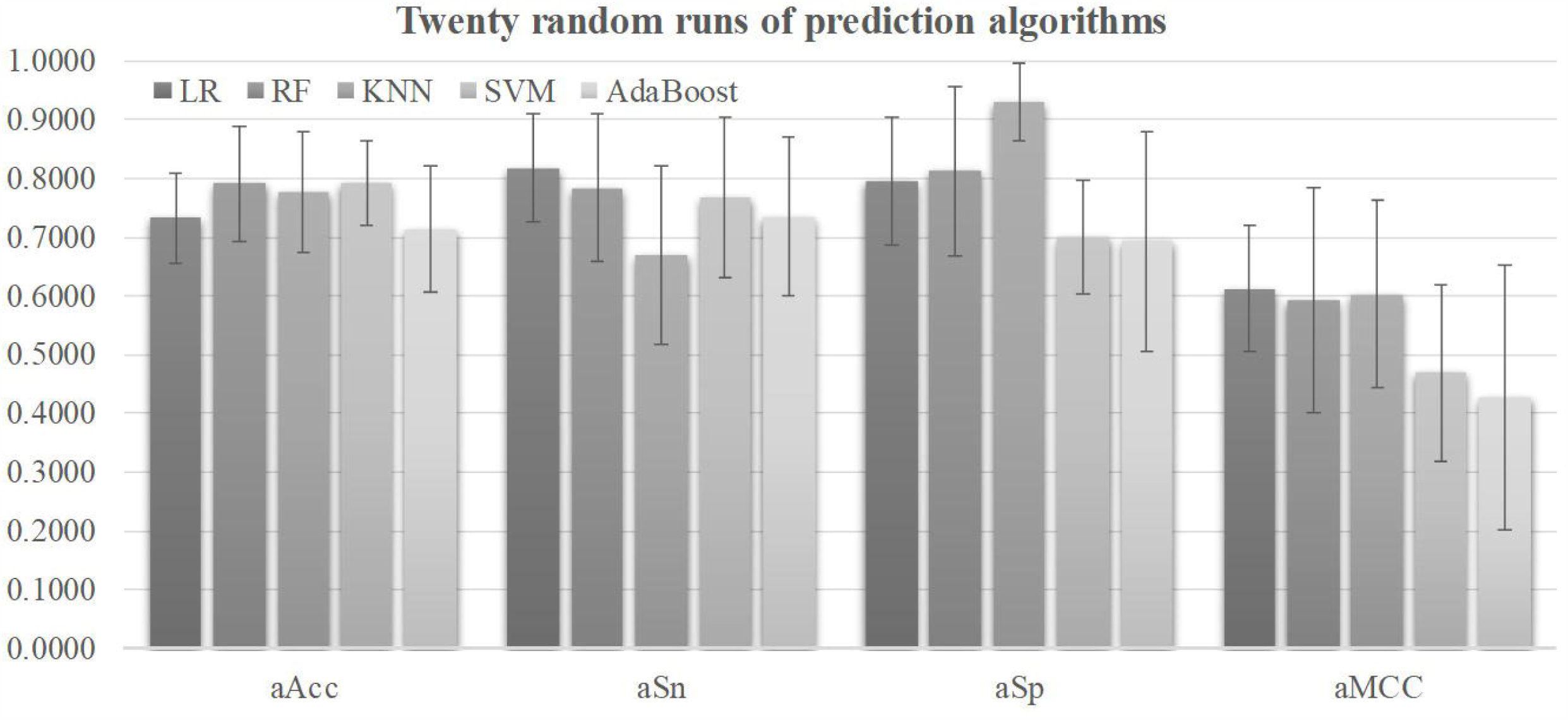
Performance metrics of five prediction algorithms. The horizontal axis was the four performance metrics, aAcc, aSn, aSp, and aMCC, which were averaged over the twenty random runs. The vertical axis gave the values of these four metrics. The bar heights and the error bars of these histograms were the averages and standard deviations of these metrics over the twenty random runs of each algorithm.

### Choosing the best threshold

A threshold may be tuned to find the balanced model performances for both positive and negative samples, as shown in Figure 3. The metric Youden’s index was introduced by W.J. Youden in 1950 to catch the best performance of a dichotomous diagnostic model [39]. Youden’s index assigns equal weights for sensitivity and specificity and tries to maximize the index value J =(Sn+Sp-1) [39]. Figure 3 illustrated the changing curves of Sn and Sp with different thresholds for the prediction scores of the samples. The maximal value of J was achieved at the threshold 0.7318, and averaged accuracy of SVM was improved to 0.8148.

**FIGURE 3.**
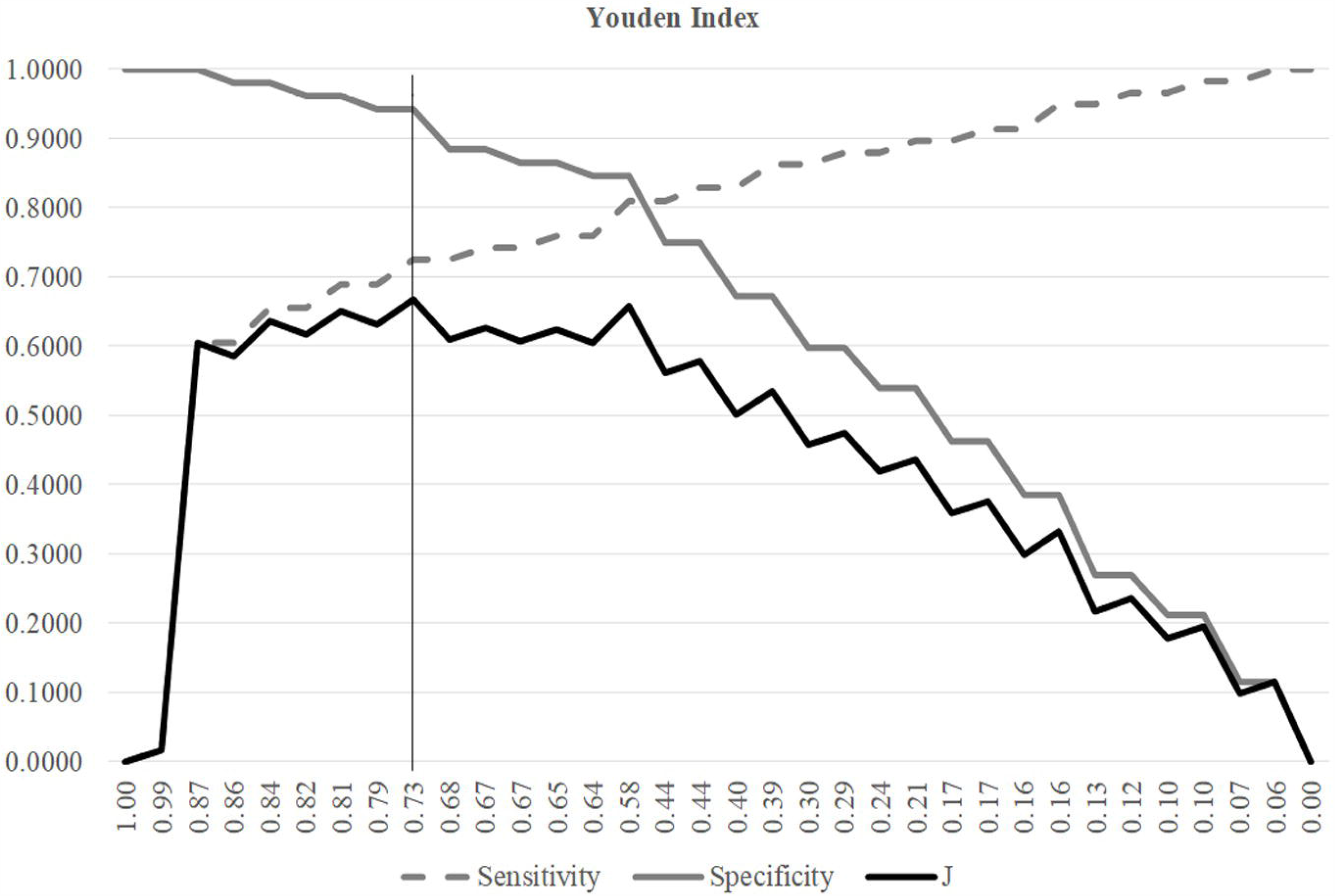
Youden index of different SVM thresholds. The three line plots were Sn, Sp and J, respectively. The horizontal axis was the threshold values sorted in the descending order. The vertical axis was the value of these three metrics Sn, Sp and J.

The Youden’s index was used to find the best threshold of the SVM models with different parameters and features in the following sections.

### Tuning the parameters of the SVM model

The grid search strategy was carried out to evaluate how different parameter values affected the disease severeness detection model, as shown in Figure 4. Parameter tuning was a time-consuming step. So this section randomly split the training dataset into 80% sub-training dataset and 20% validation dataset. Each model was trained using the sub-training dataset and the performance was calculated on the validation dataset. The model detection performance didn’t change with the linear kernel and different choices of the parameter Gamma, as shown in Figure 4 (b). And the best accuracy=0.8636 of the linear kernel SVM was achieved when C=0.1 or 1. The best model with the RBF kernel achieved Acc=0.9091 for the validation dataset, where C=100 and Gamma=0.0010. The other three metrics Sp=1.0000, Sn=0.8333 and MCC=0.8333 were also the best values in Figure 4. The SVM model with the above-mentioned parameters achieved Acc=0.8148 on the independent test dataset. So the following sections used these two choices of the parameters C and Gamma.

**FIGURE 4.**
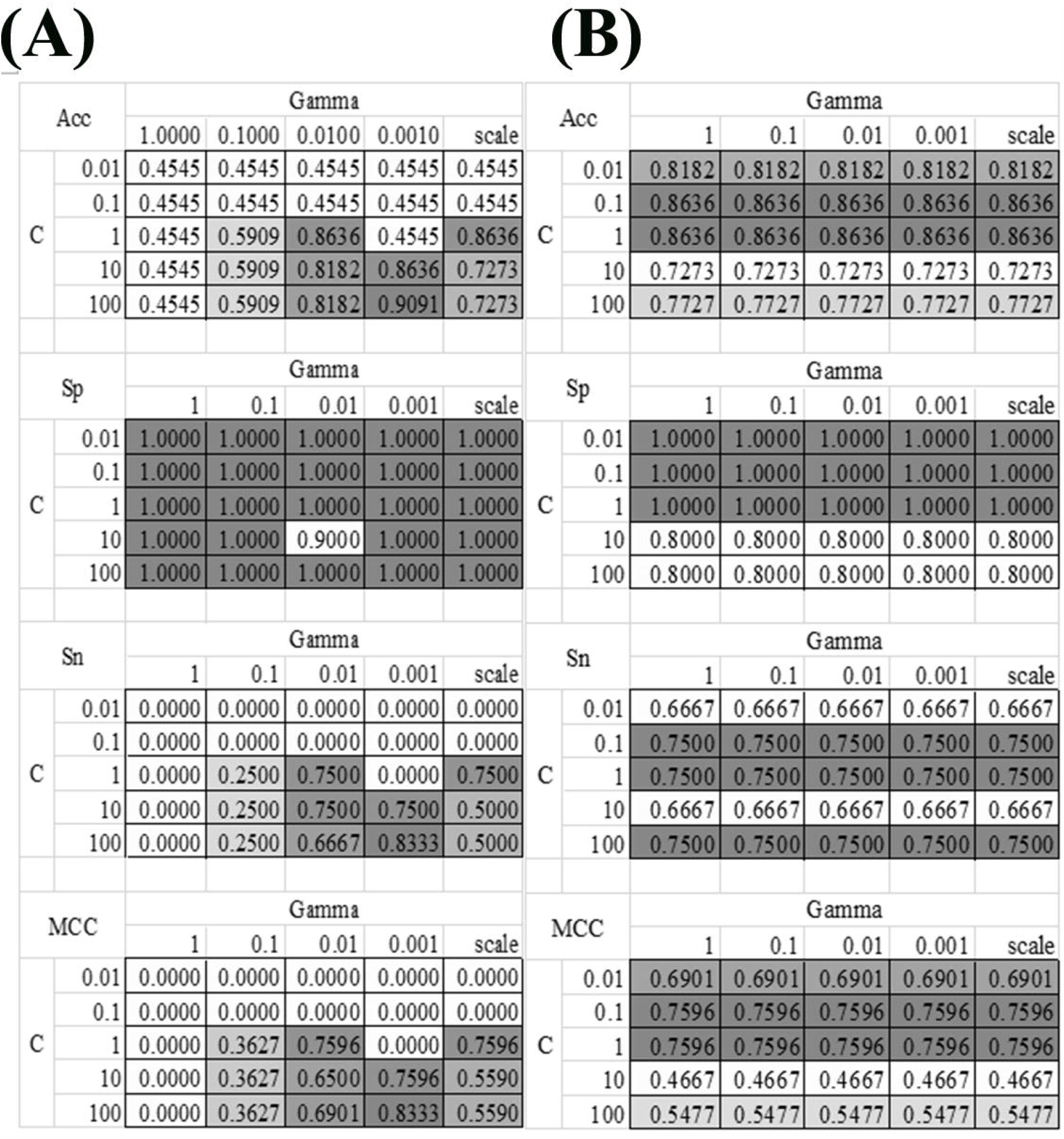
Heatmaps of the SVM parameter tuning. Two kernel functions were evaluated, i.e., **(A)** RBF and **(B)** Linear. The SVM parameter C had five value choices: 0.01, 0.1, 1, 10 and 100. The other parameter Gamma had five value choices: 1, 0.1, 0.01, 0.001 and “scale”, where “scale” was the default value in the Python library. The four detection performance metrics Acc/Sn/Sp/MCC were used to evaluate the models.

### Remove redundant features to improve the model

The existence of strong inter-feature correlations in Figure 1 (B) suggested that some features may be removed to further improve the model. This section carried out a conservative recursive feature elimination (cRFE) strategy to eliminate the redundant features while ensuring the model performance was not decreased. The model performance was evaluated for its threshold-independent metric AUC value [40,41]. Firstly, all the 32 features were ranked by the ascending order of their T-test Pvalues. Then, the detection model was evaluated by eliminating each feature. A feature was eliminated if the model’s AUC was improved with its removal. Otherwise that feature was kept. The final feature set was returned after all the features were evaluated.

The feature selection procedure should avoid using the test samples, so this section calculated the performance metrics on the validation dataset using the model trained over the sub-training dataset, as shown in Figure 5. The heuristic cRFE strategy ensured by its nature that the model performance would not be decreased, and the rising line segment indicated the removal of the feature on the horizontal axis. Four features were removed, i.e., “Age”, “Blood | Interleukin-10”, “Blood | Prothrombin time,”, and “Blood | Oxygen partial pressure”. Figure 1 (B) illustrated that all these four features were strongly correlated with some other features, with the PCC values at least 0.51.

**FIGURE 5.**
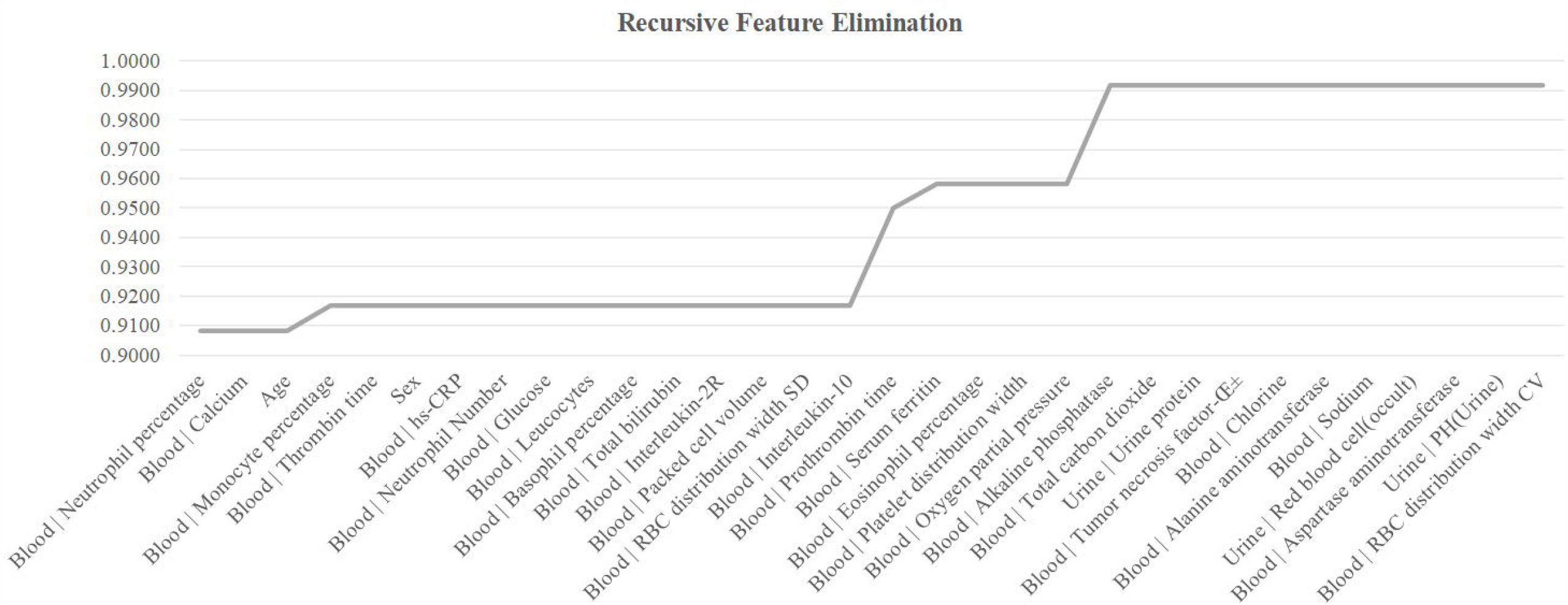
Recursive eliminating the features. The horizontal axis listed the features ranked in the ascending order by their T-test Pvalues. The vertical axis was the threshold-independent metric Area Under the Curve (AUC) achieved by the model using the feature set specified by the horizontal axis.

The remaining 28 features achieved Acc=0.9917 on the validation dataset, and Acc=0.8148 on the independent test dataset. Although the COVID-19 severeness detection performance was not improved, the model complexity was reduced and the clinical screening cost was reduced with fewer features.

A web site was established to help the clinicians to try this COVID-19 infection severity estimation model, and the users may access: http://dVirusSeverity.HealthInformaticsLab.org/.

## DISCUSSION

The emergence of SARS-CoV-2 marked the third of highly pathogenic coronavirus in humans in the twenty-first century, after severe acute respiratory syndrome (SARS) in 2003, and Middle East respiratory syndrome (MERS) in 2012 [42,43]. SARS-CoV-2 belongs to the coronavirus family, β-coronavirus genera and belongs to the cluster of betacoronaviruses [44]. Based on Sequence analysis, the amino acid sequences of SARS-CoV-2 showed 94.4% identity with SARS-CoV [45]. It is suggested that SARS-CoV-2 was more closely related to SARS-like bat CoV. In comparison, SARS-CoV-2 was more distant from the MERS-CoV [46,47]. The mortality of critically ill patients with COVID-19 is considerable. The survival time of the dead patients may be within 1-2 weeks after ICU admission [48].

The present diagnosis of COVID-19 didn’t achieve a satisfying accuracy. Both false positives and false negatives need to be decreased [49-51]. The clinical decisions of COVID-19 infections are usually confirmed by epidemiological features, clinical manifestations, imaging factors, and nucleic acid screenings, etc. Some of the COVID-19 patients may develop severe symptoms and these patients are at a much higher mortality rate than the other patients. This challenge raised the scientific question of finding the COVID-19 severeness specific biomarkers, which may help reduce the overall mortality.

This study investigated the binary classification problem between 75 severely illed COVID-19 infected patients and the other 62 patients with mild symptoms. A comprehensive optimization procedure led to the best SVM-based COVID-19 severeness detection model using only 28 features. The experimental data suggested that the severely illed patients had a higher serum level of neutrophil percentage and lower serum levels of monocyte percentage and calcium compared with those mild ones. Urine test contributed three weak group-specific biomarkers, i.e., urine pH value, urine protein and urine red blood cell. Compared with the urine pH value, the variations of urine protein and urine red blood cell were very large and these two urine features may not serve well as COVID-19 infection severeness biomarkers. The blood test features demonstrated much more significant inter-group differences than the urine test features. The summary data suggested these three blood test features as candidate severeness biomarkers, i.e., serum ferritin, hs-CRP, interleukin-2R, and tumor necrosis factor-α.

COVID-19 severeness detection model achieved the overall accuracy 0.8148 on the independent test dataset with only 28 clinical biomarkers. Twenty-one out of these 28 biomarkers were investigated in the coronavirus. Two serum values “Blood | Tumor necrosis factor-α” (56 papers) and “Blood | Sodium” (57 papers) were known to be associated with the coronavirus infections. The tumor necrosis factor-alpha (TNF-alpha) was observed to have elevated expression levels in the serum of the coronavirus-infected mice [52]. The serum sodium level was slightly increased by 2.01% in the severely ill patients in the cohort used in this study. Hoffman, et al., proposed that the pulmonary complication were more frequently observed in the hypernatremia patients [53]. So it would be interesting to investigate the underlining mechanism of how the serum sodium may induce the COVID-19 severeness. The feature “Urine | PH(Urine)” is the pH level in the urine, and quite a few investigations observed the aberrant pH levels in the body fluid or fecal matter of the coronavirus-infected animals [54,55]. Although the urine pH level was not investigated in the coronavirus-infected animals, this may be worth of an investigation. The sex bias was also observed that coronavirus tended to infect males [56,57]. Our data suggested that males were at a higher risk to be infected by COVID-19 and to develop more severe symptoms.

An accurate severeness detection model of the patients with COVID-19 based on those features may improve the prognosis of this disease in large scale clinical practices, and reduce the incidence of COVID-19 severeness and mortality. The biomarkers used for an accurate diagnosis model of patients with COVID-19 may serve as the drug targets for this global infectious disease.

There are some limitations that should be noted. First, the number of patients with COVID-19 is relatively small, which may limit the accuracy of severeness detection model. Second, since all subjects in our study were Chinese patients with COVID-19, the results may not be applied to other ethnicities. Third, the data of this study is only the preliminary establishment of COVID-19 severeness detection model. Further studies are still needed.

This study utilized the machine learning algorithms to detect the COVID-19 severely ill patients from those with only mild symptoms. Our experimental data demonstrated strong correlations with the COVID-19 severeness. And the final COVID-19 severeness detection model achieved the accuracy 0.8148 on the independent test dataset using only 28 clinical biomarkers. The detection model itself is in urgent need for the current epidemic situation that the severely ill patients are at a very high mortality rate. The 28 biomarkers may also be investigated for their underlining mechanisms of their roles in the COVID-19 severely ill patients.

## Data Availability

Data available

## AUTHOR CONTRIBUTIONS

NZ collected the data. HY, and RZ analyzed the data and wrote the paper. MD, TX, JP, EP, JH, YZ, and XX did literature search. HX, FZ and GW conceived and designed this study. The funder of the study had no role in study design, data collection, data analysis, data interpretation, or writing of the report of this study. The corresponding author had full access to all the study data and had final responsibility for the decision to submit for publication. All authors read and approved the final manuscript.

## ACKNOWLEDGMENTS

We thank all patients.

